# Evaluating and mitigating unfairness in multimodal remote mental health assessments

**DOI:** 10.1101/2023.11.21.23298803

**Authors:** Zifan Jiang, Salman Seyedi, Emily Griner, Ahmed Abbasi, Ali Bahrami Rad, Hyeokhyen Kwon, Robert O. Cotes, Gari D. Clifford

**Affiliations:** Department of Biomedical Informatics, Emory University School of Medicine, Atlanta, GA, United States; Department of Biomedical Engineering, Emory University and Georgia Institute of Technology, Atlanta, GA, United States; Department of Psychiatry and Behavioral Sciences, Emory University School of Medicine, Atlanta, GA, United States; Department of IT, Analytics, and Operations, University of Notre Dame, Notre Dame, IN, United States

## Abstract

Research on automated mental health assessment tools has been growing in recent years, often aiming to address the subjectivity and bias that existed in the current clinical practice of the psychiatric evaluation process. Despite the substantial health and economic ramifications, the potential unfairness of those automated tools was understudied and required more attention.

In this work, we systematically evaluated the fairness level in a multimodal remote mental health dataset and an assessment system, where we compared the fairness level in race, gender, education level, and age. *Demographic parity ratio (DPR)* and *equalized odds ratio (EOR)* of classifiers using different modalities were compared, along with the F1 scores in different demographic groups. Post-training classifier threshold optimization was employed to mitigate the unfairness.

No statistically significant unfairness was found in the composition of the dataset. Varying degrees of unfairness were identified among modalities, with no single modality consistently demonstrating better fairness across all demographic variables. Post-training mitigation effectively improved both DPR and EOR metrics at the expense of a decrease in F1 scores.

Addressing and mitigating unfairness in these automated tools are essential steps in fostering trust among clinicians, gaining deeper insights into their use cases, and facilitating their appropriate utilization.

**Author summary:** In this work, we systematically explored and discussed the unfairness reporting and mitigation of automated mental health assessment tools. These tools are becoming increasingly important in mental health practice, especially with the rise of telehealth services and large language model applications. However, they often carry inherent biases. Without proper assessment and mitigation, they potentially lead to unfair treatment of certain demographic groups and significant harm. Proper unfairness reporting and mitigation of these tools is the first step to building trust among clinicians and patients and ensuring appropriate application.

Using our previously developed multimodal mental health assessment system, we evaluated the unfairness level of using various types of features of the subjects for mental health assessment, including facial expressions, acoustic features of the voice, emotions expressed through language, general language representations generated by large language models, and cardiovascular patterns detected from the face. We analyzed the system’s fairness across different demographics: race, gender, education level, and age. We found no single modality consistently fair across all demographics. While unfairness mitigation methods improved the fairness level, we found a trade-off between the performance and the fairness level, calling for broader moral discussion and investigation on the topic.

## Introduction

Mental disorders are the second most common cause of years lived with disability worldwide [1], and approximately one billion people worldwide live with at least one mental disorder, most without access to effective care [2, 3]. The adoption of telehealth has been growing rapidly for mental health services, especially since the COVID-19 pandemic, to reduce cost and improve access [4–7]. Moreover, the current state of mental health clinical practice is compromised by inherent subjectivity and potential biases in the diagnostic and evaluative procedures [8, 9]. For example, African American individuals are disproportionally diagnosed with psychotic disorders [10] using both structured and unstructured clinical interviews [11]. Self-rated mental health questionnaires such as General Anxiety Disorder-7 (GAD-7) [12], and Patient Health Questionnaire-9 (PHQ-9) [13] are also highly subjective and over-reported compared to clinical diagnoses [14].

Objective automated digital assessment tools and corresponding digital biomarkers have been widely developed to aid clinicians in addressing the access, subjectivity, and bias challenges [15]. Those tools have been evaluated in various types of mental health disorders, using single data modality [16–18] or multiple modalities [19–21], in both lab-controlled [22] and remotely collected datasets [23, 24]. Although promising results have been shown in those studies, the potential bias inherent in the proposed methodologies could impede the fair diagnosis and evaluation of the underprivileged or underrepresented groups, leading to detrimental health consequences.

Addressing the potential adverse health outcomes necessitates an examination of the origins of biases in automated systems. These biases primarily stem from three sources: the mental health condition labels used for training, the construction of the dataset, and the use of pre-trained models with inherent biases. While addressing the first issue requires collaboration with clinical experts, improving fairness in the latter two aspects can be achieved during system development. There has been a growing interest in fairness research, particularly in measuring and mitigating bias in binary classification models [25].

Building upon this, a critical aspect of fairness research is the evaluation of dataset disparities, commonly assessed through the mean difference of positive labels across various demographic groups [25].A key concept in fairness evaluation based only on model output is *demographic parity (DP)* [26], which defines fairness as the equal proportion of positive predicted or estimated labels for privileged and unprivileged groups. Another type of fairness metric defines fairness as the fair prediction/classification performance of privileged and unprivileged groups, among which *equality of odds (EO)* and *equality of opportunity* [27] are the most adopted, which gauge fairness by examining disparities in true positive rates and false positive rates across different groups. To alleviate the unfairness originating from dataset formulation, training procedures, and classifier operating point selection, a range of bias mitigation strategies have emerged. These include pre-processing techniques like re-sampling and re-weighting of training data based on labels and demographic groups [28], as well as in-training fairness regularization and post-training fairness optimization approaches [27, 29], which have gained considerable traction in recent years.

As the popularity of machine learning approaches grows in medical applications, more studies have started assessing and reducing unfairness in proposed systems [30–32]. In the last two years, A growing number of studies have also applied those approaches to the mental health domain, such as drug prescription [33], phone usage-based mental health assessment [34], electrocardiogram-based anxiety prediction [35] and arrhythmia detection [36], and electronic health record-based drug misuse and depression classification [37, 38]. However, a notable research gap exists in standardized reporting [39] and evaluating the fairness of automated mental health assessment systems that employ computer vision, linguistic, and acoustic analysis techniques. Despite their longstanding central role in real-time mental health research, focused studies on their fairness, particularly in comparing different modalities and evaluating self-rated versus clinically diagnosed labels, are markedly scant. Existing studies [40, 41] primarily concentrate on gender fairness in unimodal or bimodal classifiers, leaving a significant aspect of this research area unexplored.

To address the challenges above, we evaluated the unfairness level in a multimodal remote mental health assessment system proposed in [19] for detecting mental health conditions (MHC), which was trained on a relatively diversely constructed remote mental health interview dataset. Fairness levels of the dataset itself and fairness levels of classifiers on individual features and the combination of features from diverse modalities, including facial, vocal, linguistic, and cardiovascular, were evaluated. Then, post-training threshold adjusting was applied to mitigate the unfairness in the evaluated system, and the effects of the mitigation were measured.

Group fairness metrics were adopted in this study because of the critical nature of equitable treatment across different demographics. Historical biases and systemic inequities, notably prevalent in mental health practices, can lead to disparate outcomes and significant harm to disadvantaged groups, regardless of the overall accuracy of the system. In this study, we evaluated group fairness criteria, including DP, EO, and equal accuracies across different groups, to directly assess and mitigate the long-standing biases in mental health assessments, ensuring that all groups receive equitable care.

The main contribution of this study is twofold: (1) We have provided the first systematic evaluation of the fairness level in a multimodal remote mental health assessment system, where we compared the fairness level of different modalities and different types of features, including hand-crafted features, features from supervised pre-trained deep learning models, and embedding from self-supervised-learned transformer-based [42] foundation models. (2) We have demonstrated that the existing unfairness in the system can be substantially mitigated, with reasonable trade-offs in the overall classification performance. This was achieved utilizing post-training adjustments, highlighting a promising avenue for enhancing fairness in mental health assessments.

## Materials and methods

### Dataset

The same dataset described in our previous study [19] was used in this study. The Emory University Institutional Review Board and the Grady Research Oversight Committee granted approval for this study (IRB# 00105142). For the initial screening, interviewees were recruited for either a control group (no history of mental illness within the past 12 months) or a group currently experiencing depression. After the semi-structured interview, all final diagnoses and group categorizations were verified and finalized by the overseeing psychiatrist and clinical team.

The remote interview was divided into three parts: 1) A semi-structured interview, 2) a sociodemographic section, and 3) clinical assessments, which included clinical evaluation and self-reported ratings such as General Anxiety Disorder-7 [12] and Patient Health Questionnaire-9 [13].

Briefly, data from 73 subjects were included in the analyses. They were aged 18 − 65 and were native English speakers. All interviews were conducted remotely via Zoom’s secure, encrypted, HIPAA-compliant telehealth platform. Both Video and Audio were recorded.

Subjects were categorized into three different binary categorizations based on self-rated scales or clinicians’ diagnoses. The primary categorization we discuss in this work is based on clinical diagnoses, where they were grouped into control (*n* = 22) vs. subjects with mental health conditions (MHC, *n* = 51). The latter included subjects diagnosed with any mental health condition currently or a history of diagnosis within 12 months. The control group included the remaining subjects. The categorization details can be found in our previous study [19].

Additionally, we adopted categorization based on PHQ-9 and GAD-7 scores and considered subjects self-rated depression or anxiety when the score was higher than 10. For the primary categorization, we did not find statistically significant differences in age or years of education between the control and MHC groups using Mann-Whitney rank tests. Subjects from demographic groups that did not form significantly large groups (*n <* 5) were excluded. Specifically, subjects who self-identified as Hispanic (*n* = 2) or other (*n* = 4) were not included in race-related analyses, and subjects who identified as non-binary gender (*n* = 2) were not included in gender-related analyses.

#### Recording quality assessment

Recording qualities varied across the dataset due to the differences in the subjects’ network conditions and device capabilities. While no perceivable quality difference was found for audio recordings, there exist noticeable differences in video quality. We defined a video as “low quality” if one or more of the following issues were presented in the video: having a lower resolution than “720p”, the camera not looking directly at the face, the face not fully presented due to obstruction or movement, the lighting was too dark or too bright. Then, we analyzed whether the video quality was correlated with the gender, race, age, and education level of the subjects and the performance of the classifiers using video-derived features (described in Section Multimodal assessment of mental health conditions).

### Multimodal assessment of mental health conditions

A multimodal analysis framework was proposed and evaluated in our previous study [19]. For each audiovisual interview recording, It extracted visual, vocal, language, and remote photoplethysmography (rPPG) time series signals at the frame or segment level, summarizes those time series with statistical and temporal dynamic features at the subject level (except for text embedding from the large language model, where the model directly generated subject-level embedding), and evaluates the performance of these features in clinical diagnoses (control vs. MHC) or self-rated depression/anxiety classification tasks. The details of the processing steps and system components can be found in [19].

Facial expressions and cardiovascular features were extracted from the videos, where frames were extracted, and face and facial landmarks were detected at each frame. Then, the heart rate of the subject was estimated at each frame (25 Hz) from rPPG using the *pyVHR* package [43, 44]. The probability of the presence of seven basic emotions (neutral, happiness, sadness, surprise, fear, disgust, and anger) and 12 facial action units [45] (AU1, 2, 4, 6, 7, 10, 12, 14, 15, 17, 23, 24) was estimated using convolutional neural network (CNN) based models [46, 47]. The fairness of the visual foundation model described in our previous study [19] was not evaluated due to its poor performance.

Linguistic and acoustic features were derived from the patient-side audio during the semi-structured interview. Only patient-side audio during the semi-structured interview section was used to avoid using subjects’ answers to sociodemographic or clinical assessment questions. *PyAudioAnalysis* [48] package was used to extract acoustic features at each 100ms window with 50% overlap, including zero crossing rate, energy, entropy of energy, spectral centroid/spread/entropy/flux/rolloff, Mel frequency cepstral coefficients (MFCC), and 12 chroma vector and corresponding standard deviations. For linguistic features, audio files were transcribed into texts using Amazon Transcribe on HIPAA-compliant Amazon web services at Emory, following the protocol detailed in [49]. The probability of each utterance being neutral, happy, sad, surprised, fearful, disgusted, and angry was estimated using a distilled RoBERTa model [50, 51], along with the estimation of being negative or positive using another RoBERTa-based model [52]. Beyond the language sentiments features, LLAMA-65B [53], a transformer model with 65 billion parameters, was used to generate a text embedding for the entire transcripts during the semi-structured interview.

Statistics of the time series extracted above were used as subject-level features. Both average and standard deviations over time were used for lower-dimensional (*<* 100) time series, including time series of facial expressions, acoustic features, language sentiments, and estimated heart rates from rPPG.

LLAMA-65B embedding of the entire semi-interviews was directly used as subject-level features. Additionally, hidden Markov models (HMM) with four states and a Gaussian observation model were used to model the dynamics of the low dimensional time series, using *SSM* package [54]. The statistics (duration and frequency of inferred states) of the unsupervisedly learned HMMs were used as subject-level features.

We evaluated features generated from the above-described processes in three binary classification tasks described in Section Dataset, including control vs. MHC, moderately depressed (PHQ-9 scores *>* 10, *n* = 24) vs. rest (PHQ-9 scores *<*= 10, *n* = 43), and moderate GAD severity (GAD-7 scores *>* 10, n=16) vs. rest (GAD-7 scores *<*= 10, *n* = 49). Two types of multimodal late fusion were used. The first type of fusion was the majority vote of each unimodal classifier, and the second type of fusion was a weighted vote using the probability output of the unimodal classifiers as weights. Classification performances were measured by the average macro-averaged F1 score and accuracy using a 100 times repeated stratified five-fold cross-validation.

### Fairness metrics

The fairness of the dataset and classifications were evaluated. We analyzed the fairness level of both self-rated and clinically-rated labels and focused the algorithmic fairness analysis on classifying clinically rated labels.

#### Demographic and label distribution of the constructed dataset

The number of subjects from different demographic groups and the selection rates (SR) in different groups were calculated. The selection rate was defined as the percentage of samples being “positive”, meaning clinically-rated MHC or self-rated depression, or self-rated anxiety.

#### Demographic parity ratios of the classifications

Following the *DP* defined in [26], we used two definitions of the demographic parity ratio to measure the fairness level of the classifications. For sensitive demographic variable *k* with *G* different groups and a *g*\* social-economically privileged group: The first demographic parity ratio (DPR) captured overall parity between any pairs of groups and was defined as:

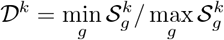

and *g* ∈ *G*_*k*_; The second demographic parity ratio focused on the parity compared to the privileged group and was defined as:

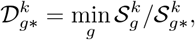

where *g* ≠ *g*\* and 𝒮 is the Selection Rate of the utilized classifier, i.e., the ratio of positive classification.

As privileged groups, we defined “male” for gender parity analysis, “white” for race parity analysis, “Older (≥ 40)” for age parity analysis, and “College or below (≤ 16 years of education)” for education parity analysis. Using classification results of the test folds in 100 repeated fold-fold cross-validation (detailed descriptions in Section Multimodal assessment of mental health conditions and [19]), DPRs of classifiers trained with features from different modalities were calculated. DPR being further from one means a larger disparity between the privileged and unprivileged groups.

### Equalized odds ratios of the classifications

Similar to DPR metrics defined in the above section, we followed *EO* definition proposed in [27] and defined the overall and over-privileged equalized odds ratios (EOR) based on false positive rate (FPR) and true positive rate (TPR). The first EOR was defined as:

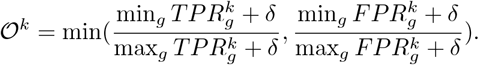

The second equalized odds ratio was defined as:

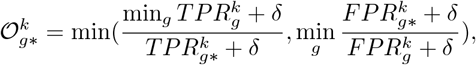

where *g* ≠ *g*\* and *δ* was set to 0.001 to avoid ratios being divided by zero. Similarly, EORs were calculated for each classifier. EOR of one means that all groups have the same TPR and FPR.Fairness metrics The fairness of the dataset and classifications were evaluated. We analyzed the fairness level of both self-rated and clinically-rated labels and focused the algorithmic fairness analysis on classifying clinically rated labels.

#### Demographic and label distribution of the constructed dataset

The number of subjects from different demographic groups and the selection rates (SR) in different groups were calculated. The selection rate was defined as the percentage of samples being “positive”, meaning clinically-rated MHC or self-rated depression, or self-rated anxiety.

#### Demographic parity ratios of the classifications

Following the *DP* defined in [26], we used two definitions of the demographic parity ratio to measure the fairness level of the classifications. For sensitive demographic variable *k* with *G* different groups and a *g*\* social-economically privileged group: The first demographic parity ratio (DPR) captured overall parity between any pairs of groups and was defined as:

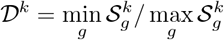

and *g* ∈ *G*_*k*_; The second demographic parity ratio focused on the parity compared to the privileged group and was defined as:

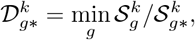

where *g* ≠ *g*\* and 𝒮 is the Selection Rate of the utilized classifier.

As privileged groups, we defined “male” for gender parity analysis, “white” for race parity analysis, “Older (≥ 40)” for age parity analysis, and “College or below (≤ 16 years of education)” for education parity analysis. Using classification results of the test folds in 100 repeated fold-fold cross-validation (detailed descriptions in Section Multimodal assessment of mental health conditions and [19]), DPRs of classifiers trained with features from different modalities were calculated. DPR being further from one means a larger disparity between the privileged and unprivileged groups.

#### Equalized odds ratios of the classifications

Similar to DPR metrics defined in the above section, we followed *EO* definition proposed in [27] and defined the overall and over-privileged equalized odds ratios (EOR) based on false positive rate (FPR) and true positive rate (TPR). The first EOR was defined as:

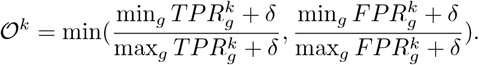

The second equalized odds ratio was defined as:

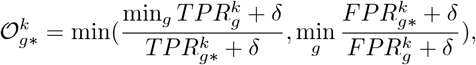

where *g* ≠ *g*\*and *δ* was set to 0.001 to avoid ratios being divided by zero. Similarly, EORs were calculated for each classifier. EOR of one means that all groups have the same TPR and FPR.

### Mitigating unfairness

For each demographic variable (gender, race, education level, age), the classifying threshold of each demographic group was learned by optimizing accuracy with equalized odds constraints, following the scoring function derived non-discriminating predictors method from Hardt et al. [27]. EORs were used as the measure for effective mitigation. Specifically, for each demographic variable, threshold adjusting was added to the cross-validation process using *Fairlearn* python package [55].

### Statistical Analyses

Mann-Whitney rank sum tests were used to assess the differences in (1) ages and years of education between controls and subjects with mental health conditions and (2) the fairness and performance of classification resulting from features from different modalities. Chi-square tests were used to assess the independence of demographic variables and video quality. Significance was assumed at a level of *p <* 0.05 for all tests.

## Results

### Fairness of the constructed dataset

The number of subjects and selection rates of being MHC from different demographic groups are shown in Fig 1. No statistically significant dependence was found between being MHC, having self-rated depression, or having self-rated anxiety and demographic variables. No statistically significant video quality differences were found either.

**Fig 1.**
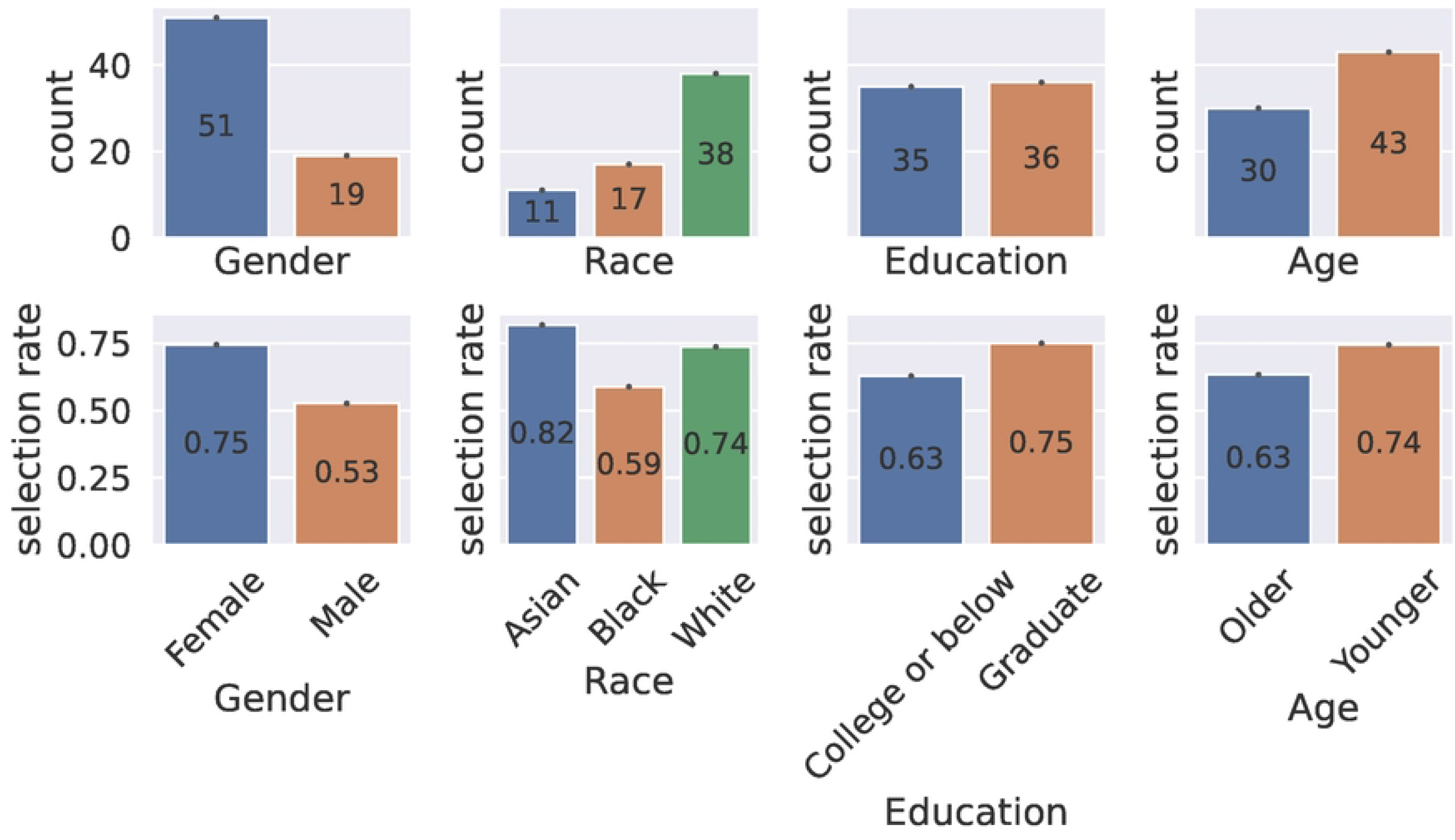
Counts and selection rates grouped by demographic groups.

### Fairness of individual modalities and in combination

Panel A in Fig 2 shows the macro-weighted F1 scores for detecting MHC using different classifiers of different demographic groups. While the number of samples available often matters to the training of the classifiers, the majority group does not always have better performance. For example, following Section Statistical Analyses, the older group significantly outperformed (*p <* 0.001) the majority younger group considering all classifiers, and the male group significantly outperformed (*p <* 0.001) the female group. Additionally, the more privileged group may not necessarily have better performance, especially when the features were less affected by the demographic variable in the analysis. For instance, the less educated group significantly outperformed (*p <* 0.001) the more educated group overall but underperformed significantly (*p <* 0.001) when the language sentiments (“Language sentiments”) were used. In contrast to education level, the privileged group in race (white group) significantly (*p <* 0.001) outperformed the underprivileged groups (Asian and Black group), while no significant differences in overall classifier performances were found between underprivileged groups.

**Fig 2.**
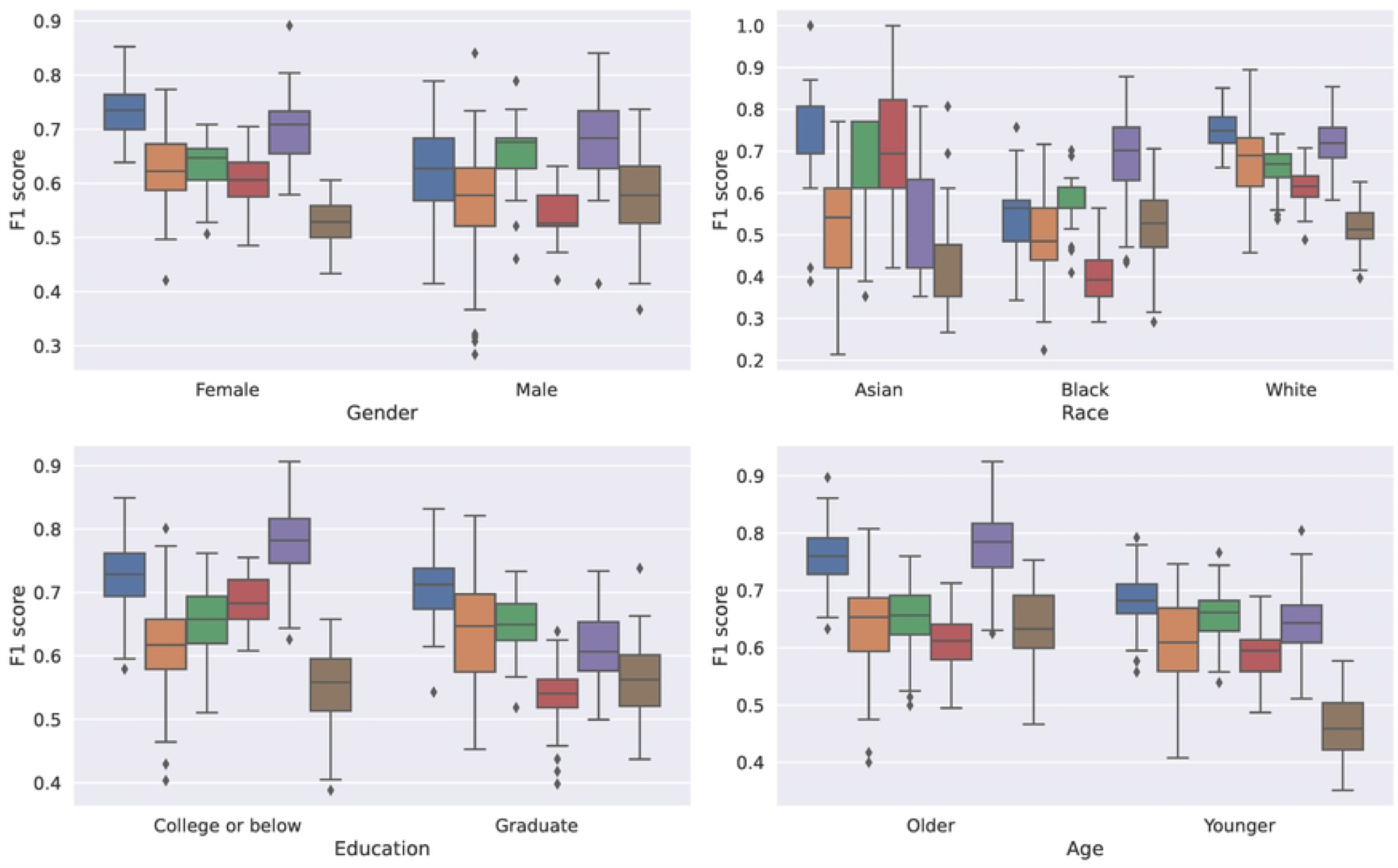
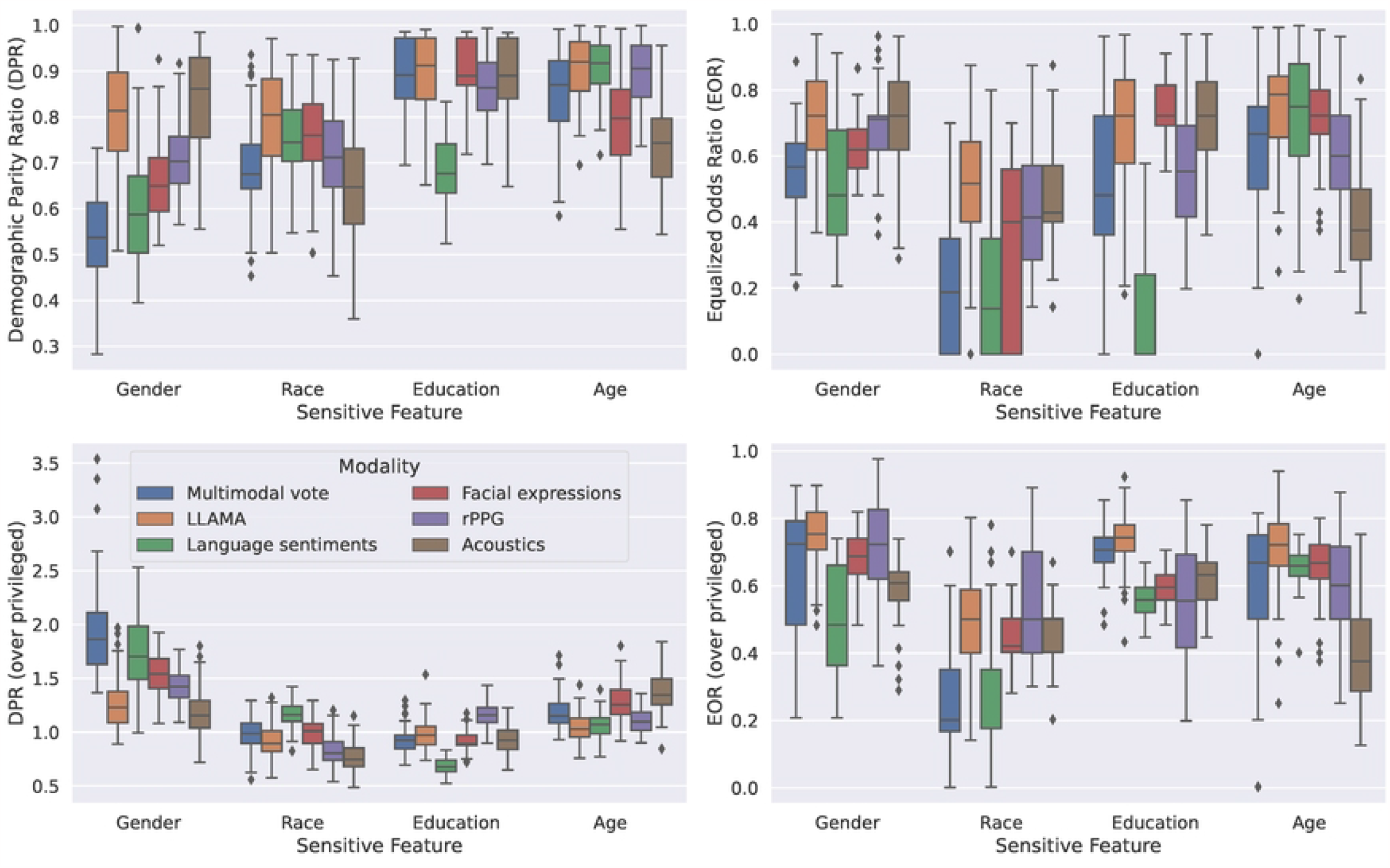
Classification performance and fairness level of different feature types in detecting MHC. (A) Classification performance by modalities. (B) Fairness by modalities. The top row shows the overall fairness metrics and the bottom row shows the fairness level when compared to the privileged group defined in Section Fairness metrics. Different colors indicate classifiers using different types of features. “Multimodal Vote” refers to the majority voting of all the unimodal classifiers. Each boxplot shows the interquartile range (IQR), where the center line shows the median value. ♦ marks outliers that are at least 1.5 IQR above the first quartile or 1.5 IQR below the third quartile.

Panel B in Fig 2 shows the fairness level (DPRs and EORs) of different classifiers in gender, race, education level, and age. It is worth noting that the fairness level of a classifier varied enormously, and the classifier that used certain types of features from one modality might have drastically different ranking among other classifiers when a different demographic variable was analyzed. Moreover, high-performing classifiers, such as multimodal voting (“Multimodal Vote”) and cardiovascular dynamics (“rPPG”), did not result in higher fairness levels. Overall, voting with probability weighting significantly (*p <* 0.001) outperformed the naive voting in both DPRs and EORs, while having just slightly lower F1 scores as shown in Fig 2. Interestingly, while large foundation models like LLAMA-65B were known to have biases toward certain demographic groups [53], classifiers based on those features showed reasonable fairness levels except for race. Using LLAMA-65B for non-white groups led to significantly lower (*p <* 0.001) F1 scores compared to the privileged group, as shown in the EOR plots and the race plots in Fig 2. Across different demographic variables, EORs and DPRs were significantly lower for race (*p <* 0.05) compared to any other demographic variables.

No significant dependence was found between video quality and the classification performances of the classifiers using video-derived facial and cardiovascular features.

### Improved fairness after mitigation

Fig 3 shows the effect of unfairness mitigation by optimizing thresholds for optimal equalized odds for different demographic variables. The results of all modalities were combined into a single box in the boxplots.

**Fig 3.**
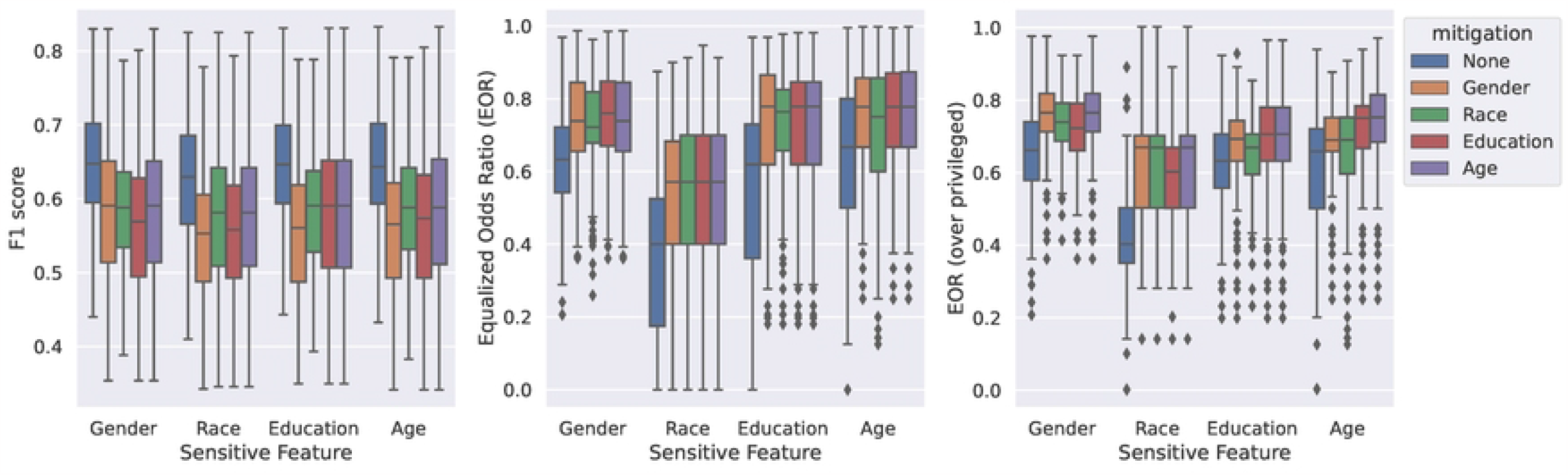
Effects of post-training mitigation on classifier performance and fairness. Different demographic variables used in the mitigation process were encoded as different colors. Similar to Fig 2, boxplots were plotted with interquartile ranges and outliers to visualize F1 scores or the EORs.

Adjusting thresholds effectively made classifiers fairer, as indicated by the increased EORs, both overall and compared to the privileged group, shown in Fig 3. Significant differences (*p <* 0.001) were found in EORs before and after mitigation. Notably, EORs increased for all demographic variables, even when only a single EO of one demographic variable was optimized. However, as a trade-off to increased EORs, F1 scores decreased by an average of 0.06 across all demographic factors and modalities (statistically significant, *p <* 0.001). Please note that no-mitigation F1 scores for different sensitive features were slightly different because not all demographic variables were available for all subjects. Please refer to Fig 1 for details.

The effect of mitigation grouped by classifier modalities can be found in S1 Fig. Increased EORs and decreased F1 scores, except for classifiers using LLAMA-65B and cardiovascular dynamics, were found.

## Discussion

### Limitations

One key limitation of this study is the relatively small number of subjects included in the analyses. This led to subjects in minority demographic groups with extremely small numbers of subjects being excluded, such as people who self-identified as non-binary gender or people who self-identified as Hispanic. While statistical tests were performed to help quantify all the comparisons made in the study, results should be interpreted accordingly. Another limitation related to the dataset is the distribution of demographic groups might not represent real-world situations. For example, while no statistically significant dependence was found between having clinician-rated or self-rated mental health conditions and being in certain demographic groups in this dataset, dependence could exist in other situations. Additionally, the subjects in the dataset were highly educated, with an average of more than 16 years of education(college graduate).

Additional fairness mitigation approaches could be applied. For example, in-training approaches such as training with multiple demographic fairness constraints [29] and adversarial training [56] could be used. Preliminary results for adding EO constraints in training following the “reduction” method proposed in [29] was found to be less effective compared to the post-training adjustment of thresholds. While the scope of this study did not include the comparison of the efficacy of different mitigation approaches, more mitigation approaches should be tested and developed to address the fairness issue in the mental health domain in the future.

### Fairness of labels and modalities

Although no significant selection rate differences were found between demographic groups in either self-rated or clinically-rated labels (shown in Section Fairness of the constructed dataset), fairness issues might arise when the number of subjects gets larger or the distribution shifts. Clinically-rated labels were known to carry bias from the clinicians due to differences in the interview process and personal experience [8], yet self-rated labels were also susceptible to subjectivity and over-reporting (higher selection rate) [14], which could be affected by demographic variables. Although this issue cannot be easily addressed, standardized reporting of the selection rate could help reduce the chance of building unfair datasets.

Furthermore, demographic variables gathered in medical applications may not comprehensively capture the physiological condition of subjects, particularly in more vaguely defined attributes such as race. Race information may carry potential biases in the data collection process when collected by clinicians and still suffer from genetic and cultural heterogeneity within commonly delineated racial groups.

Differences in fairness across modalities were partially discussed in Section Fairness of individual modalities and in combination. Many confounding variables might affect the fairness of classifiers for different demographic variables. While no significant dependence was found between video quality and the classification performances of the classifiers using video-derived facial and cardiovascular features in this dataset, the quality of the recordings could bias feature extraction models to favor certain demographic groups. For example, racial disparity was widely found in speech recognition [57], and face detection [58]. With the growing use of representation embeddings in the era of large language models and other foundation models, it is critical to understand, measure, and mitigate bias in each of the system’s building blocks.

### Future directions

With the rapid development and potential adoption of automated tools for mental health assessment in the near future, the implication of unfair evaluation and treatment could be tremendous.

#### Standardized fairness reporting

As the first barrier of unfairness in those tools, clinicians, researchers, and developers should work together to raise awareness of fairness evaluation and standardize fairness reporting practices. Understanding the fairness levels of the systems leads to the safer application of the system to minimize the risks toward underrepresented groups, even when the unfairness is not straightforward to be addressed technically [59], or when the number of subjects is relatively small.

#### Health-oriented unfairness mitigation

General fairness mitigation approaches might not fit directly into the health-related machine learning application due to the vast differences in the societal consequences due to unfair systems impacting intersections of demographic variables and health conditions. Creating unfairness mitigation methods with a clear understanding of their impact is key to the safe application of automated systems. Alday et al. [36] demonstrated that biases manifest as a function of model architecture, population, cost function, and optimization metric, all of which should be closely considered when selecting mitigation approaches. Furthermore, modality-specific debiasing methods [60] may not adapt to multi-modal health-related machine learning contexts.

#### Performance-fairness trade-off

Trade-offs between model performance and fairness should be discussed. Currently, no consensus has been made on the acceptable levels of unfairness except the U.S. Equal Employment Opportunity Commission’s “80% rule” in racial disparity presented in hiring [61]. Trade-offs in medical applications frequently demand decisions requiring broader moral discussion, especially when the overall utility cannot be clearly defined and directly measured, potentially giving rise to ethical dilemmas reminiscent of the trolley problem [62]. While specific thresholds would vary by domain and specific tasks, more research, such as investigation of the quantitative impact of unfair treatment (and of misdiagnosis), could shed light on this challenge.

## Conclusion

In this study, we rigorously assessed the fairness levels of a multimodal mental health assessment system before and after implementing unfairness mitigation strategies. These evaluations were quantitatively measured utilizing demographic parity ratios and equalized odds ratios, focusing on key demographic attributes: race, gender, education level, and age. The fairness of different modalities was compared for different demographic attributes, and post-training classifier threshold optimization successfully mitigated the unfairness with reasonable trade-offs in the overall classification performance.

## Data Availability

Data cannot be shared publicly since it contains personal identifiable information (video recordings of participants’ faces and audio recordings of speech). Although participants agreed to be recorded for purposes of the work, there are restrictions in place for the release of PII from Emory IRB. Additionally, due to the sensitive nature of the data that we collect, Certificates of Confidentiality are in place to prohibit disclosure of identifying information. Data are available from the Emory department of biomedical informatics (please contact the authors or) for researchers who meet the criteria for access to confidential data.

## Supporting information

**S1 Fig.**
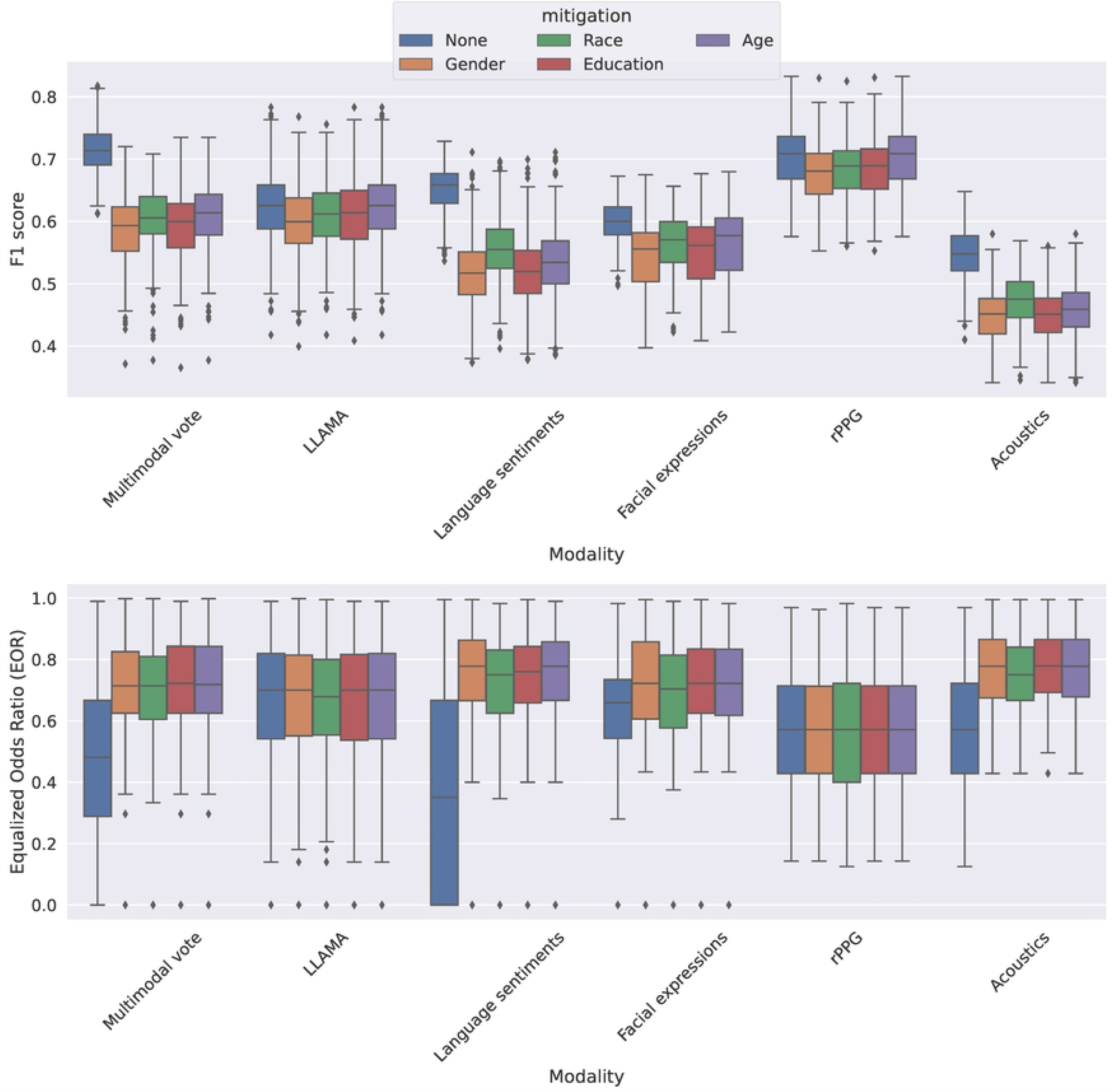
Effects of post-training unfairness mitigation on the F1 scores and EORs of classifiers using different modalities.

## Acknowledgments

The data collection described in this research was supported by funding from in part by Imagine, Innovate, and Impact Funds from the Emory School of Medicine through a Georgia Clinical & Translational Science Alliance National Institutes of Health award (UL1-TR002378). The open-access publication fee was partially supported by open access publishing fund at Emory University.

